# Risk factors for SARS-CoV-2 infection: A test-negative case-control study with additional population controls

**DOI:** 10.1101/2023.03.15.23287300

**Authors:** Marjut Sarjomaa, Chi Zhang, Yngvar Tveten, Hege Kersten, Harald Reiso, Randi Eikeland, Johny Kongerud, Kristine Karlsrud Berg, Carina Thilesen, Svein Arne Nordbø, Ingeborg S. Aaberge, Jan Paul Vandenbroucke, Neil Pearce, Anne Kristin Møller Fell

## Abstract

**Objectives:** To assess risk factors for SARS-CoV-2 infection by first comparing positive cases with negative controls as determined by polymerase chain reaction (PCR) testing and then comparing these two groups with an additional population control group.

**Design and setting:** Test-negative design (TND), multicentre case-control study with additional population controls in South Eastern Norway.

**Participants:** Adults who underwent SARS-CoV-2 PCR testing between February and December 2020. PCR-positive cases, PCR-negative controls, and additional age-matched population controls.

**Primary outcome measures:** The associations between various risk factors based on self-reported questionnaire and SARS-CoV-2 infection comparing PCR positive cases and PCR-negative controls. Using subgroup analysis, the risk factors were then compared with a population control group. Univariate and multivariate regression analyses were performed.

**Results:** In total, 400 SARS-CoV-2 PCR-positive cases, 719 PCR-negative controls, and 14,509 population controls were included. Male sex was associated with the risk of SARS-CoV-2 infection when PCR-positive cases were compared with PCR-negative controls (OR 1.9, 95% CI 1.4 to 2.6). Age, education level, comorbidities (asthma, diabetes, hypertension), an exercise were not associated with the risk of SARS-CoV-2 infection when PCR-positive cases were compared with PCR-negative controls. In the subgroup analysis comparing PCR-positive cases with age-matched population controls, asthma was associated with the risk of SARS-CoV-2 infection (OR 1.6, 95% CI 1.1 to 2.1). Daily or occasional smoking was negatively associated with the risk of SARS-CoV-2 infection in both analyses (OR 0.5, 95% CI 0.3 to 0.8 and OR 0.55, 95% CI 0.35, to 0.82, respectively).

**Conclusions:** Male sex was a possible risk factor, whereas smoking was negatively associated with the risk of SARS-CoV-2 infection, when comparing PCR-positive cases and PCR-negative controls. Asthma was associated with the risk of SARS-CoV-2 infection when PCR-positive cases were compared with population controls.

**ARTICLE SUMMARY:** *Strengths and limitations of this study:*

- The test-negative design (TND) was an important strength of this study. The design can reduce confounding from healthcare-seeking bias because PCR-controls are likely to have similar healthcare-seeking attitudes as PCR+ cases.
- This study mostly included non-hospitalised patients, which can improve the generalisability of the findings to the general public.
- The use of an additional control group from the general public for comparison with the findings from the test-negative controls provides further information on the similarities and differences in risk factors for COVID-19 and other respiratory tract infections.
- In the subgroup analyses, PCR+ cases and PCR- controls were compared with the population controls to assess the risk factors for those aged 18–55 years. Hence, the results may not be generalisable to patients older than 55 years.
- PCR test results, rather than symptoms, were used to categorise the participants into cases or controls, and therefore risk factors for SARS-CoV-2 infection and not COVID-19 disease were assessed.

## INTRODUCTION

Understanding the risk factors for severe acute respiratory syndrome coronavirus 2 (SARS-COV-2) infection is essential for prevention of new waves of Coronavirus Disease-19 (COVID-19), developing new vaccination strategies and in preparation for future pandemics. Various studies have explored the risk of SARS-CoV-2 infection as well as COVID-19 severity and mortality. Although several risk factors for SARS-CoV-2 infection and COVID-19 have been identified, findings have been conflicting (1-4), particularly with regard to the association between smoking status, obstructive lung diseases, including chronic obstructive pulmonary disease and asthma, and the risk of SARS-CoV-2 infection and development of COVID-19 (3, 5-12).

Diabetes mellitus is an important risk factor for disease severity and hospital mortality (1, 3, 13). In a meta-analysis of adults hospitalised in 11 countries, overweight and diabetic patients were more likely to require respiratory support (13). Another meta-analysis showed that obesity was associated with the COVID-19 susceptibility and severity (14). Hypertension has also been linked with COVID-19 severity (2). In addition, advancing age ≥ 60 years and male sex were associated with increased mortality (2). Air pollution can be a risk factor for upper and lower respiratory tract diseases. There are few previous studies that report association of concentrations of particulate pollutants in cities and COVID-19 incidence (15). To our knowledge no studies have investigated environmental factor, such as air pollution from wood-fired heating, as potential risk factor for SARS-CoV-2 infection (15).

COVID-19 varies from asymptomatic infection to mild pneumonia and may lead to serious respiratory illness and death. Most SARS-CoV-2 infected individuals have mild symptoms and are not hospitalised, but few studies assess these patients (16). To date, most studies have been retrospective, designed as traditional case-control studies and cohort studies involving hospitalised patients, and have demonstrated substantial heterogeneity among findings (1, 6, 17, 18). In contrast, our test-negative design (TND) multicentre case-control study, using two different control groups differs from the classical case-control study in that the controls are defined by a negative test result (19-21). This study design can reduce potential bias resulting from differences in health care-seeking attitude between cases and controls (19). Further, we utilize a population control group from the pre-existing Telemark general population study dataset, which included a random sample of 14,509 participants included in 2018 (22). The additional control group makes it possible to assess similarities and differences between the PCR- negative and general population control group.

Polymerase chain reaction (PCR) tests are the gold standard for detection of SARS-CoV-2 (23). Therefore, the aim of this study was to assess the risk factors for SARS-CoV-2 infection by comparing individuals with PCR positive (PCR+) and PCR negative (PCR-) tests as part of a TND case-control study with additional population controls (19-21).

## METHODS

### Study design and setting

We designed a TND case-control study with additional population controls. Participants were defined as ‘cases’ or ‘controls’ based on their PCR+ and PCR- test results, respectively. We used the first PCR test results of each participant during the inclusion period in our analysis.

First, we compared SARS-CoV-2 PCR- positive individuals (cases) with SARS-CoV-2 PCR- negative individuals (controls) in a classic TND study. The participants were recruited from the counties of Agder and Telemark in South-Eastern Norway from February to December 2020 during the first and second waves of the COVID-19 pandemic, and when the SARS-CoV-2 Alpha variant was dominant. The participants were recruited by telephone from all hospitals in the region, municipal laboratories, and COVID-19 test centres. The participants were chronologically selected from the PCR test lists during the study period. The inclusion criteria were (i) adults aged ≥ 18, (ii) SARS-CoV-2 RT-PCR test result, and (iii) resident of South-Eastern Norway, specifically Agder and Telemark counties, during the inclusion period. Participants who were unable to answer the questionnaire, which was conducted in Norwegian, were excluded.

The official Norwegian testing criteria for SARS-CoV-2 changed over time but were the same for the PCR+ and PCR- participants in the study period. In the first wave of the pandemic, PCR testing was restricted to symptomatic patients. In the second wave, PCR testing was additionally applied to close contacts and asymptomatic individuals during the outbreaks. Participants were included regardless of their symptoms. Only 58 participants (5%) in the PCR+ cases and PCR- controls were asymptomatic. Most PCR+ cases had mild symptoms, with only 22 (6%) participants hospitalised during the study period (24). We aimed to include 400 PCR+ cases, and two PCR- controls matched for test time of administration and geographical location per PCR+ case to increase the power of the study.

Next, we compared PCR+ cases and PCR- controls (aged 18–55 years) with an age-matched population control group (aged 21–55 years) as part of a subgroup analysis. The population control group data were obtained from the pre-existing Telemark study dataset which included a random sample of 14,509 participants, aged 21 to 55, residing in Telemark, Norway in 2018 (22). Given that this dataset was collected more than 1 year before the pandemic, it was assumed to be PCR- negative for SARS-CoV-2. By adding population controls and using the TND, we compared PCR+ cases with two different control groups.

We used STROBE case-control reporting guidelines for our study (25).

### Questionnaire design

We used questions from the Norwegian Health Institute COVID-19 questionnaire and the Telemark study questionnaire (22, 26, 27), in addition a few questions were provided by the study group. The questionnaire consisted of questions related to (1) education status, (2) smoking habits, (3) respiratory symptoms and/or diseases, such as asthma or chronic obstructive pulmonary disease (COPD), (4) comorbidities, (5) exercise, and (6) environmental exposure to pollution from traffic or wood-fired heating. Questions are shown in the Supplementary **table S1**.Self-reported questionnaire data from the Telemark study dataset were used as population controls in the extended TND (22).

### Statistical analysis

The mean, standard deviation (SD), and median were reported for continuous variables, as appropriate. Categorical data were reported as frequencies and percentages. Logistic regression models were used to assess the possible risk factors. Univariate analysis was performed for each factor after adjusting for age and sex. Multivariate analysis was performed using all of the predictors. To determine the association between risk factors and PCR test positivity, odds ratios (OR) were reported with 95% confidence intervals (CI). Statistical analyses were performed using R (version 4.2; R Core Team, Vienna, Austria).

Our questionnaire had a low rate of missing data, ranging from 0% to 6.2% for each question, with the exception of those related to smoking habits, which had 11.2% and 15.3% missing data for PCR+ and PCR- participants, respectively. Due to the subsequent follow-up questions, the questionnaire used in the population control group had no missing data for questions related to smoking habits, asthma, COPD, diabetes, hypertension, and wood heating. For the same reason, there were no missing for the question about wood heating in our study. The remaining questions among the population controls had a low rate of missing data, ranging from 1.8% to 6.2%, with the exception of exercise which was 13.5% missing. We did not perform data imputation. We assumed that the data had values missing at random.

### Patient and public involvement

According to the Norwegian National Guidelines for User Involvement in Health Research in May 2018, two user representatives of SARS-CoV-2 PCR- positive patients were involved. They played an active role in all project phases, including the development and testing of questionnaires. The user representatives helped us understand the patient perspective, gave feedback on protocols, study methods, information and consent forms, and questionnaires, and participated actively in the dissemination of results achieved until now. All study results are also communicated via www.sthf.no/helsefaglig/forskning-og-innovasjon/forskningsprosjekter/covita and www.sshf.no/helsefaglig/forskning-og-innovasjon/covita-studien.

## RESULTS

Of 656 eligible PCR+ participants and 923 eligible PCR- participants, 400 PCR + cases and 719 PCR- controls were included. The study flow chart is shown in **Figure 1**. The characteristics and comorbidities of the PCR+, PCR-, and population controls are shown in **Table 1**.

**Table 1.**
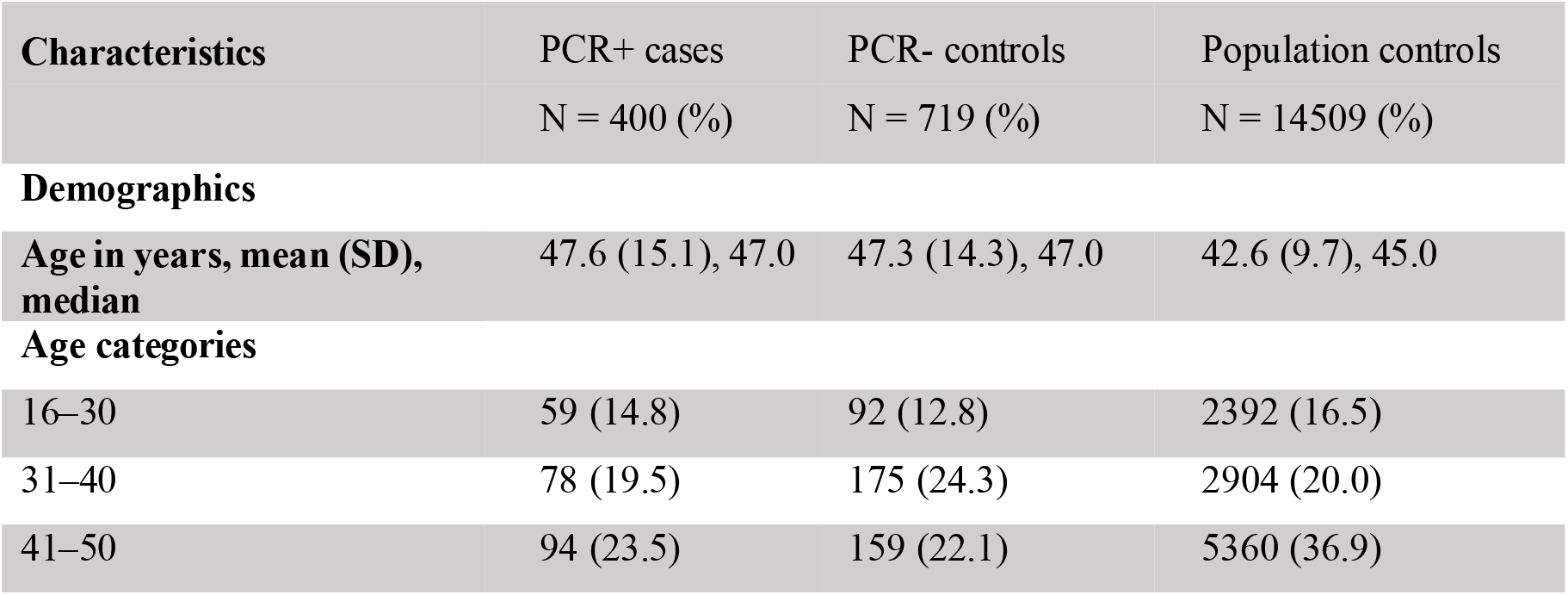
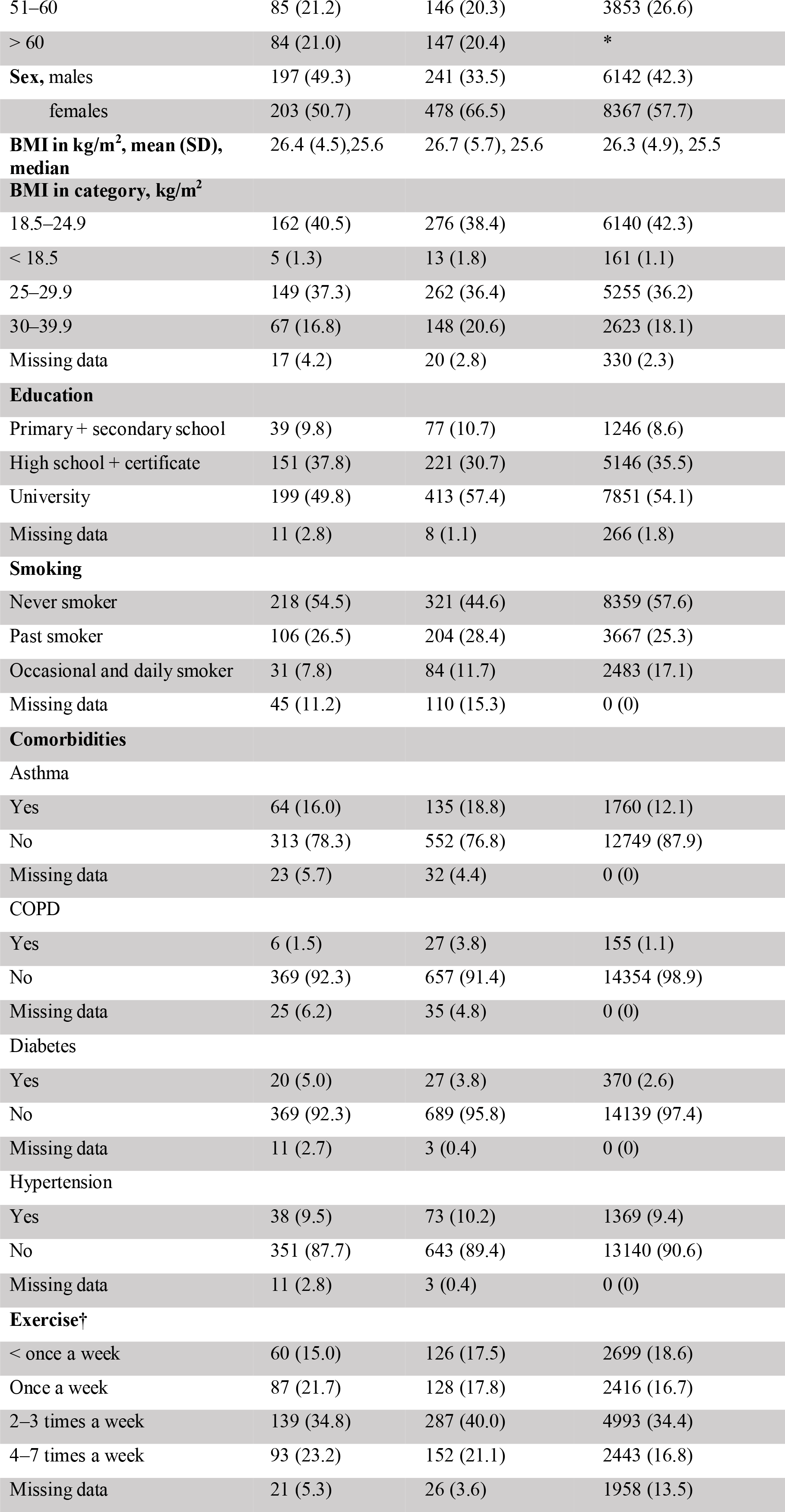
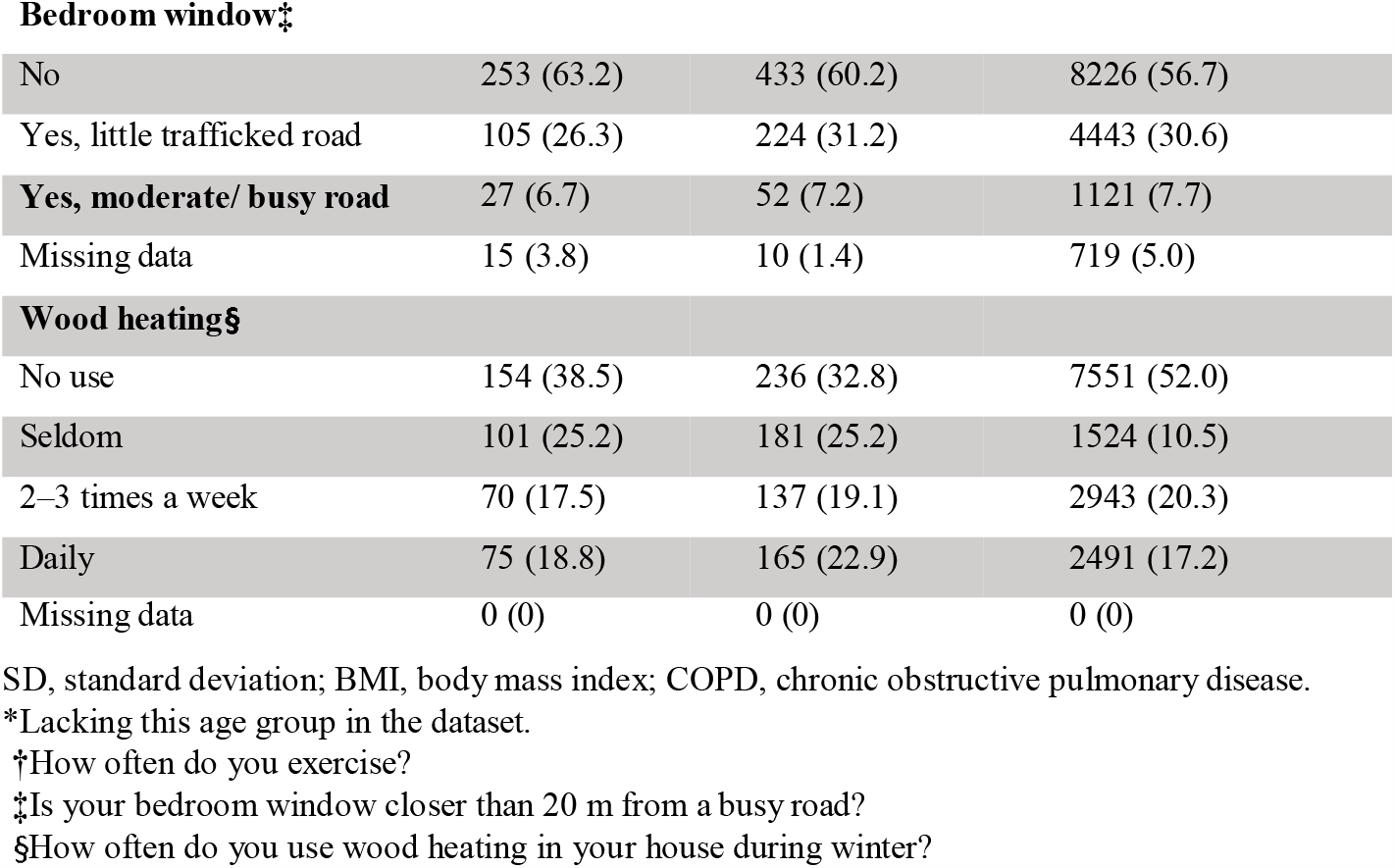
Characteristics of the PCR+ cases and PCR- controls 3–5 months after PCR test and the population control group data from the pre-existing Telemark study dataset.

**Figure 1.**
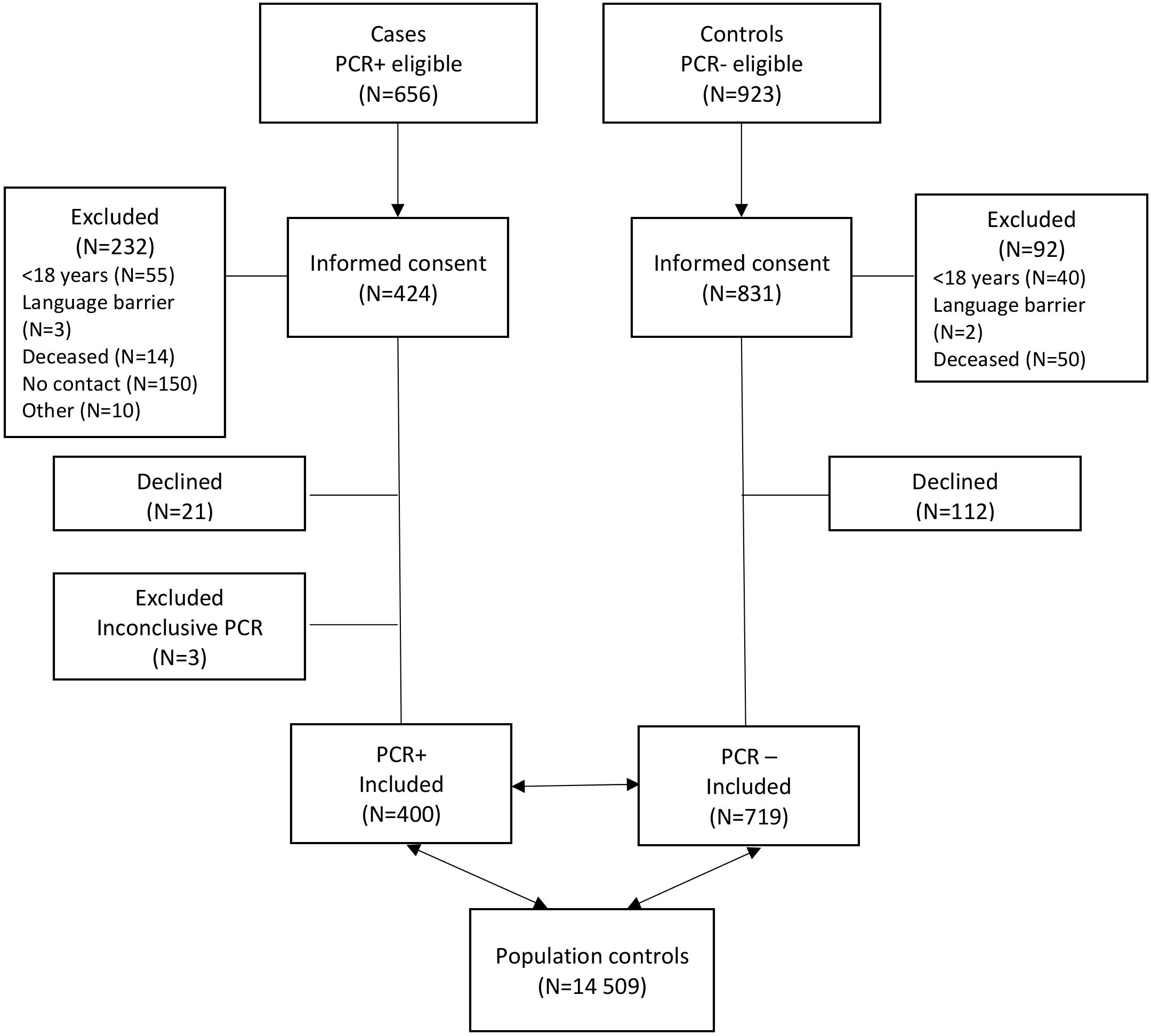
Flow chart for inclusion of participants in this study.

The PCR+ cases and PCR- controls had a mean age of 48±15 years and 47±14 years, respectively. Male participants represented 49% PCR+, 34% PCR-, and 42% of the population controls. Asthma was present in 64 (16.0%) of the PCR+ group, 135 (18.8%) of the PCR- group, and 1,760 (12.1%) of the population controls.

Characteristics and comorbidities for the subgroup analysis of the PCR+ cases and PCR- controls and population control group are shown in Supplementary **Table S2**.

The univariate and multivariate regression analyses for possible SARS-CoV-2 infection risk factors for the PCR+ cases and PCR- controls are presented in **Table 2**.

**Table 2.**
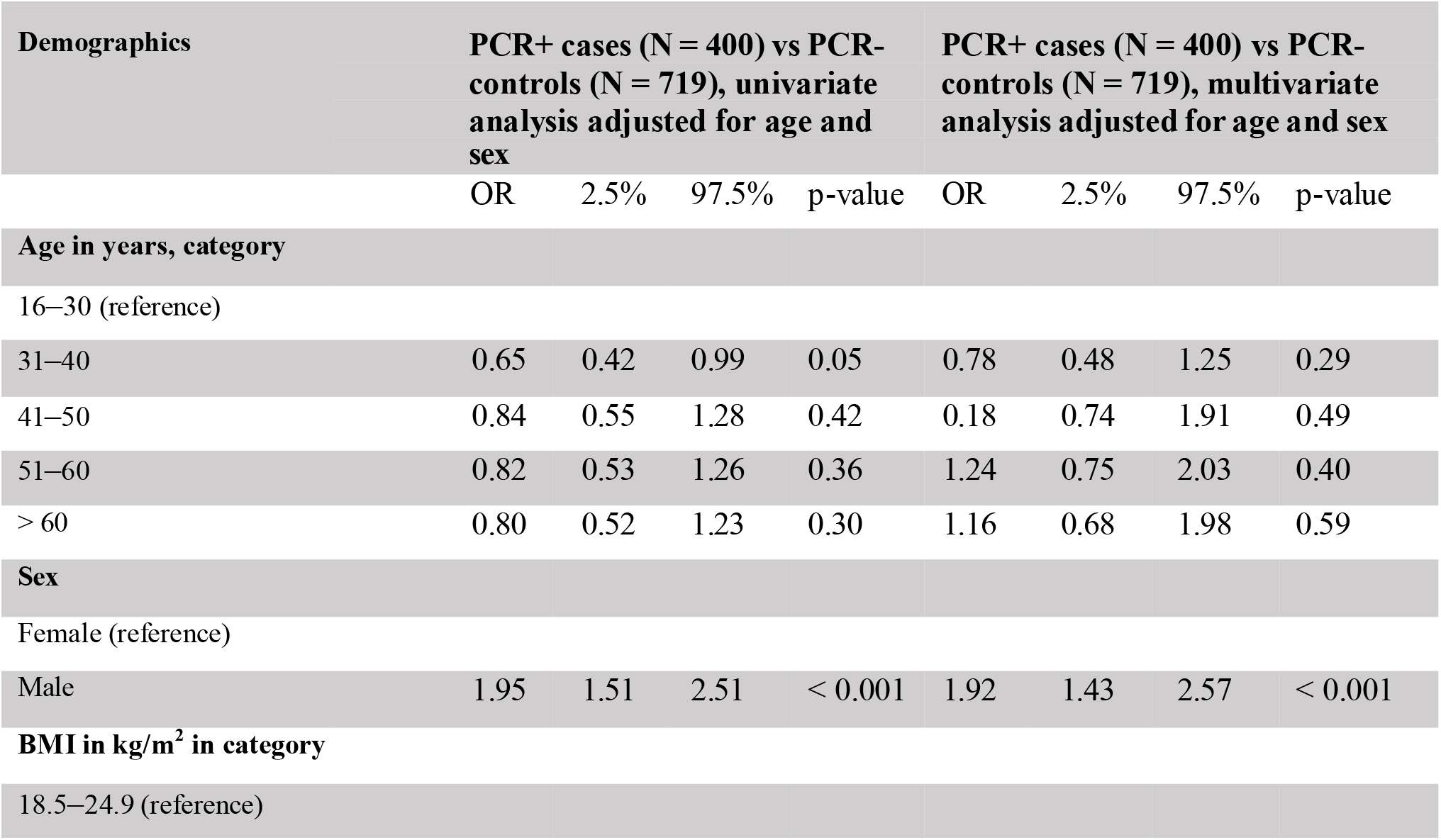
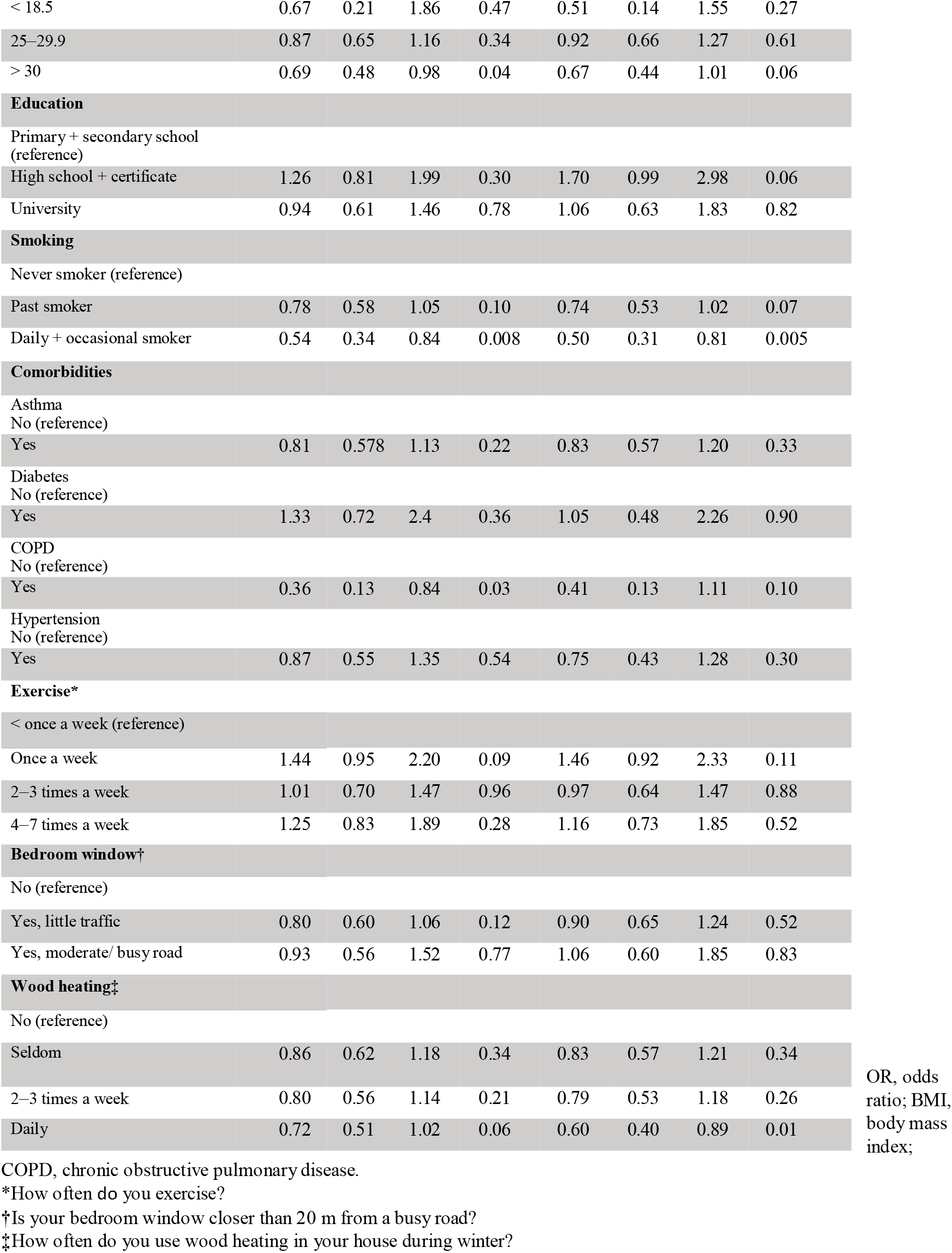
Univariate and multivariate regression analyses for possible SARS-CoV-2 infection risk factors for PCR+ cases vs PCR- controls adjusted for age and sex.

Male sex was significantly associated with the risk for SARS-CoV-2 infection when comparing PCR+ cases and PCR- controls (OR 1.92, 95% CI 1.43 to 2.57; p < 0.001). Age, education level, and comorbidities were not associated with SARS-CoV-2 infection. Daily or occasional smoking was negatively associated with SARS-CoV-2 infection (OR 0.50, 95% CI 0.31 to 0.81; p = 0.005).

The subgroup analysis of participants aged 18–55 years comparing PCR+ cases with two different control groups, PCR- controls and population controls are shown in **Table 3**.

**Table 3.** Multivariate regression analysis for SARS-CoV-2 infection risk factors for a subgroup of PCR+ cases and PCR- controls and the same subgroup of PCR+ cases and population controls adjusted for age and sex.

|  |  |  |  |  |  |  |  |  |
| --- | --- | --- | --- | --- | --- | --- | --- | --- |
| Demographics | PCR+ cases (N = 286) vs PCR-controls (N = 502), multivariate analysis adjusted for age and sex (Aged 18–55) |  |  |  | PCR+ cases (N = 286) vs population controls (N = 14509), multivariate analysis adjusted for age and sex (Aged 18–55)* |  |  |  |
| Age in years and category | OR | 2.5% | 97.5% | p-value | OR | 2.5% | 97.5% | p-value |
| **18–30 (reference)** |  |  |  |  |  |  |  |  |
| 31–40 | 0.78 | 0.48 | 1.26 | 0.31 | 1.03 | 0.71 | 1.50 | 0.89 |
| 41–50 | 1.22 | 0.76 | 1.98 | 0.42 | 0.67 | 0.46 | 0.97 | 0.03 |
| 51–55 | 1.29 | 0.78 | 2.14 | 0.32 | 0.92 | 0.63 | 1.35 | 0.67 |
| **Sex** |  |  |  |  |  |  |  |  |
| **Female (reference)** |  |  |  |  |  |  |  |  |
| Male | 1.77 | 1.28 | 2.14 | 0.32 | 1.20 | 0.93 | 1.54 | 0.16 |
| **BMI in kg/m<sup>2</sup> in category** |  |  |  |  |  |  |  |  |
| **18.5–24.9 (reference)** |  |  |  |  |  |  |  |  |
| < 18.5 | 0.69 | 0.18 | 2.23 | 0.56 | 1.65 | 0.50 | 4.10 | 0.34 |
| 25–29.9 | 0.96 | 0.67 | 1.37 | 0.80 | 1.06 | 0.80 | 1.41 | 0.66 |
| > 30 | 0.68 | 0.43 | 1.06 | 0.09 | 1.12 | 0.77 | 1.59 | 0.55 |
| **Education** |  |  |  |  |  |  |  |  |
| **Primary + secondary school (reference)** |  |  |  |  |  |  |  |  |
| High school + certificate | 1.93 | 1.03 | 3.75 | 0.05 | 1.14 | 0.69 | 1.20 | 0.62 |
| University | 1.12 | 0.61 | 2.15 | 0.72 | 1.00 | 0.60 | 1.70 | 0.93 |
| **Smoking** |  |  |  |  |  |  |  |  |
| **Never smoker (reference)** |  |  |  |  |  |  |  |  |
| Past smoker | 0.64 | 0.44 | 0.93 | 0.02 | 0.88 | 0.65 | 1.17 | 0.39 |
| Daily + occasional smoker | 0.48 | 0.28 | 0.79 | 0.004 | 0.55 | 0.35 | 0.82 | 0.005 |
| **Comorbidities** |  |  |  |  |  |  |  |  |
| **Asthma (no is reference)** |  |  |  |  |  |  |  |  |
| Yes | 0.83 | 0.56 | 1.25 | 0.38 | 1.56 | 1.12 | 2.14 | 0.007 |
| **Diabetes (no is reference)** |  |  |  |  |  |  |  |  |
| Yes | 1.00 | 0.36 | 2.61 | 0.99 | 1.28 | 0.57 | 2.49 | 0.51 |
| **COPD (no is reference)** |  |  |  |  |  |  |  |  |
| Yes | 0.45 | 0.06 | 2.04 | 0.34 | 0.77 | 0.13 | 2.49 | 0.72 |
| **Hypertension (no is reference)** |  |  |  |  |  |  |  |  |
| Yes | 0.71 | 0.32 | 1.51 | 0.39 | 0.37 | 0.19 | 0.65 | 0.001 |
| **Exercise (< once a week is reference)†** |  |  |  |  |  |  |  |  |
| Once a week | 1.68 | 1.01 | 2.81 | 0.05 | 2.02 | 1.35 | 3.05 | < 0.001 |
| 2–3 times a week | 1.28 | 0.80 | 2.05 | 0.31 | 1.47 | 1.01 | 2.19 | 0.05 |
| 4–7 times a week | 1.33 | 0.80 | 2.25 | 0.28 | 1.85 | 1.23 | 2.83 | 0.004 |
| **Bedroom window (no is reference)‡** |  |  |  |  |  |  |  |  |
| **Yes, little traffic** | 1.02 | 0.73 | 1.44 | 0.89 | 0.94 | 0.72 | 1.23 | 0.67 |
| **Yes, moderate/busy road** | 1.14 | 0.60 | 2.13 | 0.68 | 0.77 | 0.46 | 1.22 | 0.30 |
| **Wood heating (no is reference)§** |  |  |  |  |  |  |  |  |
| **Seldom** | 0.77 | 0.50 | 1.17 | 0.22 | 4.25 | 3.07 | 5.86 | < 0.001 |
| **2–3 times a week** | 0.79 | 0.51 | 1.23 | 0.30 | 1.65 | 1.17 | 2.30 | 0.003 |
| **Daily** | 0.61 | 0.39 | 0.94 | 0.03 | 2.13 | 1.50 | 2.99 | < 0.001 |

OD, odds ratio; BMI, body mass index; COPD, chronic obstructive lung disease.
\* The population control group was aged 21–55 years.
†How often do you exercise?
‡Is your bedroom window nearer than 20 m from the trafficked road?
§How often do you use wood heating in your house during winter?

Comparison of PCR+ cases with population controls in the subgroup analysis revealed that exercising once a week (OR 2.02, 95% CI 1.35 to 3.05), 2–3 times a week (OR 1.47, 95% CI 1.01 to 2.19), and 4–7 days a week (OR 1.85, 95% CI 1.23 to 2.83), having asthma (OR 1.56, 95% CI 1.12 to 2.14) and using wood heating seldom (OR 4.25, 95% CI 3.07 to 5.86), 2_3 times a week (OR 1.65, 95% CI 1.17 to 2.30) and daily during the winter (OR 2.13, 95% CI 1.50 to 2.99) were associated with SARS-CoV-2 infection. Comparison of PCR+ cases with PCR- controls or with the population controls revealed that daily or occasional smoking (OR 0.48, 95% CI 0.28 to 0.79) and (OR 0.55, 95% CI 0.35 to 0.82), respectively, was negatively associated with SARS-CoV-2 infection. Hypertension was negatively associated with SARS-CoV-2 infection when PCR+ cases were compared with population controls (OR 0.37, 0.19 to 0.65,). Age, body mass index (BMI), education level, and comorbidities were not associated with SARS-CoV-2 infection when comparing PCR+ and PCR- controls.

The regression analysis comparing the PCR- and population control groups is shown in Supplementary **Table S3**.

## DISCUSSION

In this TND study, we identified the male sex as a risk factor for SARS-CoV-2 infection in patients. In addition, smoking was negatively associated with SARS-CoV-2 infection in both the TND and population control analyses. Asthma demonstrated a non-statistically significant negative association with SARS-CoV-2 infection in the TND and a positive association when comparing PCR+ cases with the population controls. COPD showed a negative association with SARS-CoV-2 infection in both control group analysis. While exercising and wood heating during the winter were highlighted as possible risk factors for SARS-CoV-2 infection when comparing PCR+ cases with population controls, this was not the case when comparing with PCR- participants.

Male sex was associated with SARS-CoV-2 infection, which is in line with previous studies (28). In a systematic review and meta-analysis, a higher ratio of COVID-19 in males than females (100:82.5) was reported (16, 28). A meta-analysis also showed a higher risk for SARS-CoV-2 infection in men than that in women, with a relative risk of 1.08 (16).

In our study, current smoking status was negatively associated with SARS-CoV-2 infection. This paradoxical finding is reflected in the literature, with many studies reporting discordant results (5, 12, 29, 30). The lack of severe COVID-19 symptoms among our participants can have contributed to this result, with only 6% of our participants hospitalised. Moreover, the PCR- controls in our study had other common respiratory infections, which may be associated with smoking (12). Previous studies have shown that current smokers have worse outcomes associated with COVID-19 (8, 29, 30). Given that we did not obtain data related to the pack-years or duration of smoking, our results should be interpreted with caution.

Asthma and COPD were not associated with SARS-CoV-2 infection in the TND, which is comparable to studies from the early phase of the pandemic (5, 9). This may be due to the willingness of individuals with asthma to be tested whenever they develop respiratory symptoms that could indicate COVID-19. In a TND, the PCR- controls will have signs and symptoms of respiratory tract infections other than COVID-19. Thus, asthma may still be associated with COVID-19, as well as with other respiratory tract infections. A large cohort study from the United Kingdom reported that those with well-controlled mild asthma did not have a significantly increased risk of hospitalisation or COVID-19 related mortality than healthy individuals (11). Interestingly, in our study, asthma was associated with SARS-CoV-2 infection in the subgroup analysis comparing PCR+ cases with population controls. The reason for this finding is not clear; however, we observed a relatively high prevalence of asthma (16%) among the PCR+ cases in our study, but not COPD (1.5%). In many COVID-19 studies, a low prevalence of asthma (1%) and varying prevalence of COPD (2-14%) for SARS-CoV-2 infected patients have been reported (5-7, 9). It is possible that patients with chronic diseases may have isolated more than others during lockdowns in some countries or regions. The potential protective immunity provided by therapies used to treat chronic respiratory diseases may also explain the low prevalence of COVID-19 among adults with asthma or COPD in some studies (5, 7, 9). On the contrary, more severe asthma and prescription of medium- or high-dose inhaled corticosteroids have been considered as risk factors for COVID-19 (11).

In a study by Lacedonia et al., the prevalence of COPD and current smokers was low, but when infected with SARS-CoV-2, these groups had the highest all-cause mortality (5). A nationwide Korean study showed that COPD was associated with an increased risk of COVID-19 susceptibility; however, the prevalence of COPD among severe COVID-19 patients or COVID-19 mortality did not increase, but smoking influenced COPD outcomes (10). The heterogeneity of these findings may be attributed to study design, such as the selection of controls, sample size, and geographical location.

Our study demonstrated no association between age, BMI, education level, diabetes, having a bedroom window closer a trafficked road and SARS-CoV-2 infection. Hypertension was inversely associated with SARS-CoV-2 infection, which contradicts other studies (7). Obesity has been associated with COVID-19 susceptibility and severity (17, 31), and is thought to be an important prognostic factor (4, 14, 17, 31, 32). Diabetes has also been proposed as a risk factor for developing severe COVID-19 and mortality (1, 3, 13). Given that our study mostly included patients with mild COVID-19 symptoms and few hospitalised participants, this may have contributed to the finding of no associations between these factors and SARS-CoV-2 infection.

In the subgroup analysis comparing PCR+ cases and population controls, asthma, exercise and wood heating were possible risk factors for SARS-CoV-2 infection. However, given the possibility of selection bias due to differences in socioeconomic status or health care-seeking attitudes, these findings should be interpreted with caution. Analysing PCR+ cases with PCR- participants as controls in TND may have reduced this bias.

We obtained different results for asthma, exercise, and wood heating in the TND analysis than in the subgroup analyses using population controls, although findings for sex, age, smoking, and COPD showed similar directions of association. However, the interpretation of how smoking habits affect the risk of SARS-CoV-2 infection requires further assessment owing to the limited study size.

This study had some limitations. First, the questionnaire was conducted in South-Eastern Norway during a period when the SARS-CoV-2 Alpha variant was dominant; therefore, these results may not be entirely representative of other countries or virus strains. However, Telemark and Agder have both rural and urban areas and are considered to well-represent Nordic populations. Second, we compared PCR+ cases and PCR- controls with population controls to assess the risk factors for individuals aged 18–55 years; hence, the results of our subgroup analyses may not be generalisable among those > 55 years.

Third, there is a possibility of recall bias due to the use of a self-reported questionnaire; however, questions included were comparable to other studies (22, 33), including studies assessing COVID-19 (26, 27). Fourth, confounding unknown factors are possible in all epidemiological studies. Theoretically, misclassification of controls in TND may be more likely than in classical case-control studies (21). Misclassification of cases was considered less likely due to the high sensitivity of PCR tests (34, 35). We confirmed also a high specificity of the PCR tests with only few PCR- controls with positive antibodies in our previous study (24).

With the TND, which is often used for vaccine studies, it is possible to identify risk factors that are specific for COVID-19 by adding population controls (21, 36, 37). Furthermore, in the traditional TND design, participants are included before the test results. In our study, all individuals that matched our inclusion criteria were recruited and defined as cases or controls regardless of symptoms and depending only on their PCR test results (38, 39). However, we did not consider this as a limitation because the majority of the participants in our study had symptoms.

Healthcare-seeking attitude as a possible source of selection bias may be reduced with TND, as both groups have the same reason for testing (40, 41). In contrast to traditional case-control studies, controls are tested for the disease under study and are those with negative test results without exception. Although the criteria for PCR testing changed, the differences in the groups due to variation of testing strategies during the two pandemic phases were reduced because the PCR tests were matched for time and place. To assess if the PCR- control group accurately represented the source population, an additional general population control group was utilised. As demonstrated in our study, the choice of test-negative or population controls can effect outcomes regarding risk factors for SARS-CoV-2 infection.

Overall, selecting appropriate study designs and combining all relevant information from studies assessing risk factors for SARS-CoV-2 infection and COVID-19 are vital for the prevention of new waves of COVID-19 and other pandemics in the future. In particular, findings from TND studies assessing risk factors may also contribute to the development of new vaccination strategies. However, further research is needed to address the evolution of virus variants, uptake of vaccination, and differences in humoral and cellular protective immunity among risk groups.

## CONCLUSION

In this study, there were differences in the risk factors for SARS-CoV-2 infection between the test-negative design and when using population controls. Male sex was associated with the risk of SARS-CoV-2 infection when comparing PCR+ cases and PCR- controls, while smoking was negatively associated with SARS-CoV-2 infection in both the test-negative design and when using population controls. This discordance may partly be explained by the differences in healthcare-seeking attitudes among the PCR+ group compared to the population control group.

## Data Availability

There are legal and ethical restrictions on sharing our dataset. Our dataset is not fully anonymized and has a relative small sample size making identification
possible. The potentially identifying patient information is age, birthdate, location and dates for PCR testes. However, data requests for the minimal dataset, which includes only the main variables of the final analyses, can be made to the Research department at the Telemark Hospital trust, Ulefossvegen 55, 3710 Skien, Norway.

## Acknowledgements

The authors would like to thank Trude Belseth Sanden, Astrid Bjørkeid, June Bakstevold, Gølin Finkenhagen Gundersen, Emile van Gelderen, Elin Skjørvold Christensen, and Louise Myrland, Signe Seljåsen, Mona Brekke, Siv Stigen, Anne Cecilie Tveiten, Oda Eikeland Myrnes, and Siri Cathrine Rølland for their assistance with data collection and analysis. The authors would also like to express their gratitude to all participants and user representatives involved in this study. We would like to thank Editage (www.editage.com) for English language editing.

## Author Contributions

AKMF, JVP and NP were involved in the conception and design. MS, RE, KKB, HR, CT and YT contributed to data collection. CZ was responsible for the statistical analysis. All authors were involved in the interpretation of the results and contributed to manuscript writing. AKMF, YT, HK, NP, JVP and JK provided advice.

## Patient consent for publication

Not required.

## Ethics approval

All participants provided written informed consent before inclusion. The Regional Committee for Medical and Health Research Ethics of Southeast Norway A (ID 146469), Norwegian Centre for Research Data (ID 533954), and data protection officers in the participating hospitals approved the study (ID 20-02553 and ID 20-06971).

## Data availability statement

There are legal and ethical restrictions on sharing our dataset. Our dataset is not fully anonymized and has a relative small sample size making identification possible. The potentially identifying patient information is age, birthdate, location and dates for PCR testes. However, data requests for the minimal dataset, which includes only the main variables of the final analyses, can be made to the Research department at the Telemark Hospital trust, Ulefossvegen 55, 3710 Skien, Norway.

## Supplemental material

