## Supplemental table 1 for "Risk factors for SARS-CoV-2 infection: A test-negative case-control study with additional population controls"

**Supplementary table S1: Questionnaire**

| **Questionnaire** | | | | |
|---|---|---|---|---|
| **Education** | | | | |
| What is your highest education? | Primary + secondary school | High school + certificate? | University | |
| **Smoking habits** | | | | |
| Do you smoke? | Yes | No | If yes, daily? | If yes, occasionally? |
| Are you a past smoker? | Yes | No | | |
| **Comorbidities** | | | | |
| Asthma and chronic obstructive pulmonary disease (COPD) | Has a physician ever diagnosed you with asthma? | Has a physician ever diagnosed you with COPD? | | |
| Do you have the following diseases? | Diabetes | High blood pressure | | |
| **Exercise** | | | | |
| How often do you exercise? | Once a week | 2-4 times a week | 4-7 times a week | |
| **Environmental exposure** | | | | |
| Is your bedroom window nearer than 20 meter from a busy road | Yes, little trafficked road | Yes, moderate/busy trafficked road | | |
| **Wood heating** | | | | |
| How often do you use wood heating in your house during winter? | No use | Seldom | 2-3 times a week | Daily |
