## Supplemental Table 2 for "Risk factors for SARS-CoV-2 infection: A test-negative case-control study with additional population controls"

**Table S2** Subgroup analysis of cases and controls (aged 18–55 years) and population control group (aged 21–55 years). Characteristics and comorbidities for the PCR+ cases, PCR- controls at the baseline 3–5 months after PCR test and the population control group data from pre-existing Telemark study dataset from 2018**.**

| **Characteristics** | PCR+ cases | PCR- controls | Population controls |
|---|---|---|---|
| | N=286 (%) | N=502 | N=14509 |
| **Demographics** | Ages 18–55 years | Ages 18–55 years | Ages 21–55 years |
| **Age in years, mean (SD), median** | 40.2 (9.8), 41.0 | 39.8 (9.2), 40.0 | 42.6 (9.7), 45.0 |
| **Age categories** | | | |
| **16–30** | 59 (20.6) | 92 (18.3) | 2392 (16.5) |
| **31–40** | 78 (27.3) | 175 (34.9) | 2904 (20.0) |
| **41–50** | 94 (32.9) | 159 (31.7) | 5360 (36.9) |
| **51–60** | 55 (19.2) | 76 (15.1) | 3853 (26.6) |
| **> 60** | * | * | * |
| **Sex, males** | 132 (46.2) | 162 (32.3) | 6142 (42.3) |
| **females** | 154 (53.8) | 340 (67.7) | 8367 (57.7) |
| **BMI in kg/m$^2$, mean (SD), median** | 26.2 (4.6), 25.5 | 26.7 (5.8), 25.5 | 26.3 (4.9), 25.6 |
| **BMI category, kg/m$^2$** | | | |
| **18.5–24.9** | 118 (41.3) | 198 (39.4) | 6140 (42.3) |
| **< 18.5** | 4 (1.4) | 10 (2.0) | 161 (1.1) |
| **25–29.9** | 109 (38.1) | 176 (35.1) | 5255 (36.2) |
| **30–39.9** | 45 (15.7) | 105 (20.9) | 2623 (18.1) |
| **Missing data** | 10 (3.5) | 13 (2.6) | 330 (2.3) |
| **Education** | | | |
| **Primary + secondary school** | 22 (7.7) | 46 (9.2) | 1246 (8.6) |
| **High school + certificate** | 104 (36.4) | 143 (28.5) | 5146 (35.5) |
| **University** | 152 (53.1) | 307 (61.2) | 7851 (54.1) |
| **Missing data** | 8 (2.8) | 6 (1.29) | 266 (1.8) |
| **Smoking** | | | |
| **Never smoker** | 173 (60.5) | 249 (49.6) | 8359 (57.6) |
| **Past smoker** | 64 (22.4) | 134 (26.7) | 3667 (25.3) |
| **Occasional and daily smoker** | 25 (8.7) | 66 (13.1) | 2483 (17.1) |
| **Missing data** | 24 (8.4) | 53 (10.6) | 0 (0) |
| **Comorbidities** | | | |
| **Asthma** | | | |
| **Yes** | 46 (16.1) | 94 (18.7) | 1760 (12.1) |
| **No** | 225 (78.7) | 389 (77.5) | 12749 (87.9) |
| **Missing data** | 15 (5.2) | 19 (3.8) | 0 (0) |
| **COPD** | | | |
| **Yes** | 2 (0.7) | 7 (1.4) | 155 (1.1) |
| **No** | 269 (94.1) | 476 (94.8) | 14354 (98.9) |
| **Missing data** | 15 (5.2) | 19 (3.8) | 0 (0) |
| **Diabetes** | | | |

| | | | |
|---|---|---|---|
| **Yes** | 9 (3.1) | 13 (2.6) | 370 (2.6) |
| **No** | 269 (94.1) | 486 (96.8) | 14139 (97.4) |
| **Missing data** | 8 (2.8) | 3 (0.6) | 0 (0) |
| **Hypertension** | | | |
| **Yes** | 12 (4.2) | 21 (4.2) | 1369 (9.4) |
| **No** | 266 (93.0) | 478 (95.2) | 13140 (90.6) |
| **Missing data** | 8 (2.8) | 3 (0.6) | 0 (0) |
| **Exercise†** | | | |
| **< once a week** | 40 (14.0) | 98 (19.5) | 2699 (18.6) |
| **Once a week** | 66 (23.1) | 97 (19.3) | 2416 (16.7) |
| **2–3 times a week** | 102 (35.7) | 189 (37.6) | 4993 (34.4) |
| **4–7 times a week** | 64 (22.4) | 103 (20.5) | 2443 (16.8) |
| **Missing data** | 14 (4.9) | 15 (3.0) | 1958 (13.5) |
| **Bedroom window‡** | | | |
| **No** | 169 (59.1) | 297 (59.2) | 8226 (56.7) |
| **Yes, little traffic** | 85 (29.7) | 157 (31.3) | 4443 (30.6) |
| **Yes, moderate/busy road** | 21 (7.3) | 40 (8.0) | 1121 (7.7) |
| **Missing data** | 11 (3.8) | 8 (1.6) | 719 (5.0) |
| **Wood heating in the season§** | | | |
| **No** | 104 (36.4) | 156 (31.1) | 7551 (52.0) |
| **Seldom** | 70 (24.5) | 124 (24.7) | 1524 (10.5) |
| **2–3 times a week** | 56 (19.6) | 103 (20.5) | 2943 (20.3) |
| **4-7 times a week** | 56 (19.6) | 119 (23.7) | 2491 (17.2) |
| **Missing data** | 0 (0) | 0 (0) | 0 (0) |

BMI, body mass index; COPD, chronic obstructive pulmonary disease.
*Data not available.
†How often do you exercise?
‡Is your bedroom window closer than 20 m from a busy road?
§Do you have wood heating in your house during winter?
