## Supplemental Table 3 for "Risk factors for SARS-CoV-2 infection: A test-negative case-control study with additional population controls"

**Table S3** The univariate and multivariate regression analysis for a subgroup of PCR- controls vs population control group from the Telemark study. Age groups 18–55 years are included for PCR- controls and 21–55 years for the population control group.

| **Demographics** | **PCR- controls vs population controls, univariate analysis adjusted for age and sex** (Aged 18–55) | | | | **PCR- controls vs population controls, multivariate analysis adjusted for age and sex** (Aged 18–55) | | | |
|---|---|---|---|---|---|---|---|---|
| | OR | 2.5% | 97.5% | p-value | OR | 2.5% | 97.5% | p-value |
| Age categories | | | | | | | | |
| 16–30 (reference) | | | | | 1.22 | 0.92 | 1.62 | 0.17 |
| 31–40 | 1.58 | 1.22 | 2.05 | < 0.001 | 0.50 | 0.37 | 0.67 | < 0.001 |
| 41–50 | 0.79 | 0.61 | 1.02 | 0.07 | 0.67 | 0.50 | 0.92 | 0.006 |
| 51–60 | 1.01 | 0.78 | 1.33 | 0.92 | * | | | |
| > 60 | * | | | | | | | |
| Sex | | | | | | | | |
| Female (reference) | | | | | | | | |
| Male | 0.67 | 0.56 | 0.80 | <0.001 | 0.62 | 0.51 | 0.76 | < 0.001 |
| BMI in kg/m$^2$ in category | | | | | | | | |
| 18.5–24.9 (reference) | | | | | | | | |
| < 18.5 | 1.48 | 0.72 | 2.70 | 0.24 | 2.02 | 0.96 | 3.82 | 0.04 |
| 25–29.9 | 1.22 | 1.00 | 1.49 | 0.05 | 1.17 | 0.94 | 1.46 | 0.16 |
| > 30 | 1.40 | 1.11 | 1.76 | 0.004 | 1.51 | 1.16 | 1.96 | 0.002 |
| **Education** | | | | | | | | |
| Primary + secondary school (reference) | 1 | | | | | | | |
| High school + certificate | 0.71 | 0.53 | 0.98 | 0.03 | 0.61 | 0.423 | 0.87 | 0.006 |
| University | 0.85 | 0.64 | 1.15 | 0.27 | 0.78 | 0.56 | 1.11 | 0.16 |
| **Smoking** | | | | | | | | |
| Never smoker (reference) | **1** | | | | | | | |
| Past smoker | 1.43 | 1.17 | 1.76 | < 0.001 | 1.40 | 1.13 | 1.74 | 0.002 |
| Daily + occasional smoker | 0.97 | 0.74 | 1.26 | 0.84 | 1.07 | 0.80 | 1.40 | 0.66 |
| **Comorbidities** | | | | | | | | |
| Asthma No (reference) | 1 | | | | | | | |
| Yes | 0.977 | 0.811 | 1.174 | 0.804 | 0.815 | 0.555 | 1.162 | 0.277 |
| Diabetes No (reference) | 1 | | | | | | | |
| Yes | 1.10 | 0.61 | 1.82 | 0.74 | 1.25 | 0.68 | 2.13 | 0.44 |
| COPD No (reference) | 1 | | | | | | | |
| Yes | 1.97 | 0.96 | 3.6 | 0.04 | 1.43 | 0.59 | 2.95 | 0.38 |
| Hypertension No (reference) | 1 | | | | | | | |
| Yes | 0.71 | 0.49 | 1.00 | 0.06 | 0.52 | 0.34 | 0.78 | 0.002 |
| **Exercise†** | | | | | | | | |
| Never (reference) | 1 | | | | | | | |
| Once a week | 1.11 | 0.85 | 1.46 | 0.44 | 1.15 | 0.85 | 1.55 | 0.37 |
| 2–3 times a week | 1.08 | 0.85 | 1.37 | 0.55 | 1.12 | 0.87 | 1.47 | 0.38 |
| 4–7 times a week | 1.22 | 0.93 | 1.60 | 0.15 | 1.33 | 0.99 | 1.80 | 0.061 |
| **Bedroom window‡** | | | | | 1.324 | 0.543 | 2.751 | 0.491 |

| | | | | | | | | |
|---|---|---|---|---|---|---|---|---|
| No (reference) | 1 | | | | | | | |
| Yes, little traffic | 0.98 | 0.81 | 1.17 | 0.80 | 0.97 | 0.79 | 1.18 | 0.75 |
| Yes, moderate/busy road | 0.89 | 0.63 | 1.21 | 0.47 | 0.526 | 0.342 | 0.778 | 0.002 |
| **Wood heating in the season§** | | | | | | | | |
| No (reference) | 1 | | | | | | | |
| Seldom | 4.00 | 3.17 | 5.03 | < 0.001 | 1.68 | 1.31 | 2.13 | < 0.001 |
| 2–3 times a week | 1.67 | 1.31 | 2.12 | < 0.001 | 2.00 | 1.53 | 2.60 | < 0.001 |
| Daily | 2.49 | 1.98 | 3.13 | < 0.001 | 3.55 | 2.75 | 4.57 | < 0.001 |

OR, odds ratio; BMI, body mass index; COPD, chronic obstructive pulmonary disease.
*Data not available.
†How often do you exercise?
‡Is your bedroom window closer than 20 m from a busy road?
§How often do you use wood heating in your house during winter?
